# Inference-based time-resolved cortical stability and chaos analysis for focal epileptic seizures

**DOI:** 10.1101/2024.11.27.24317815

**Authors:** Yun Zhao, David B. Grayden, Mario Boley, Yueyang Liu, Philippa J. Karoly, Mark J. Cook, Levin Kuhlmann

## Abstract

Epilepsy affects millions globally, with a significant subset of patients suffering from drug-resistant focal seizures. Understanding the underlying neurodynamics of seizure initiation and propagation is crucial for advancing treatment and diagnostics. In this study, we present a novel, inference-based approach for analyzing the temporal evolution of cortical stability and chaos during focal epileptic seizures. Utilizing a multi-region neural mass model, we estimate time-varying synaptic connectivity from intracranial electroencephalography (iEEG) data collected from individuals with drug-resistant focal epilepsy. Our analysis reveals distinct preictal and ictal phases characterized by shifts in cortical stability, heightened chaos in the ictal phase, and highlight the critical role of inter-regional communication in driving chaotic cortical behaviour. We demonstrate that cortical dynamics are consistently destabilized prior to seizure onset, with a transient reduction in instability at seizure onset, followed by a significant increase throughout the seizure. This work provides new insights into the mechanisms of seizure generation and offers potential biomarkers for predicting seizure events. Our findings pave the way for innovative therapeutic strategies targeting cortical stability and chaos to manage epilepsy.

## 1. Introduction

Epilepsy, characterized by recurrent unprovoked seizures, affects approximately 50 million people worldwide and remains a significant challenge [1]. Despite advancements in neuroimaging and pharmacotherapy, a substantial proportion of individuals with epilepsy continue to experience drug-resistant seizures [2, 3, 4]. Understanding the neurophysiological mechanisms of epileptic seizures is crucial for developing new diagnosis methods and effective treatments.

Epileptic seizures are disturbances of the normal electrical activity of the brain [5]. These disturbances are often manifested as sudden, excessive discharges of neuronal activity [6]. Many studies have recognized the pathology of epileptic seizures as an interplay between *γ*-aminobutyric acid (GABA) mediated inhibition, glutamate mediated excitation, and neuronal synchrony [7, 8, 9, 10]. Crucial findings have highlighted that an imbalance between excitatory and inhibitory neurotransmitters can lead to hyperexcitability and hypersynchronous activity in neuronal networks [11, 12]. Neurophysiological mechanisms underlying these seizures involve alterations in ion channel function, receptor activity, and synaptic plasticity. For instance, mutations in ion channels, such as sodium and potassium channels, have been implicated in increasing neuronal excitability [13, 14]. Additionally, changes in GABAergic inhibition, either through altered receptor function or neurotransmitter availability, contribute to reduced inhibitory control, exacerbating seizure susceptibility [15]. Furthermore, studies have shown that network-level synchronization plays a vital role in seizure propagation, where neurons in epileptogenic regions (regions where focal seizures originate) become highly synchronized, leading to the spread of epileptic activity across the brain [16, 17].

Model-based studies have significantly advanced our understanding of epileptic seizures. Network-level models have been used to simulate the electrical activity of neuronal networks, providing insights into the dynamics of seizure initiation and propagation [18, 19, 20]. Local models help elucidate the role of various regional factors, such as synaptic connectivity, neuronal excitability, and membrane potentials, in generating seizures [21, 22, 23].

Despite these advancements, there is hitherto only a few investigations exploring seizure dynamics of networked cerebro-cortical brain systems, defined in the context of dynamical systems theory. This leads to one of the rarely investigated areas of seizure study: examining how cortical stability and chaos vary during different phases of seizures. Although some studies have described the progression of chaos in cortical activity during epileptic seizures by calculating the maximum Lyapunov exponent from intracranial electroencephalogram (iEEG) time series [24, 25, 26], the lack of corresponding neurophysiological models has hampered the interpretation of the relationship between chaos and underlying biological processes. Neurophysiological models can also help investigate how perturbations alter the stability of brain networks [27, 28].

The use of neural mass models, which simulate the collective behavior of neuron populations [29], has provided insights into the complex interactions within neural networks during seizures [30, 21, 31, 32, 33] but more so in a local modelling context. In this study, we leveraged a multi-region neural mass model to investigate the chaos and stability of focal epileptic seizures in a inference-based time-resolved way across numerous cortical sites. Model parameters and states were estimated from intracranial electroencephalography (iEEG) data of individuals with drug-resistant focal epilepsy using the NeuroProcImager tool [34]. The dynamic stability and chaotic behavior were subsequently calculated based on these parameter estimates. Across a vast number of 3008 seizures, our findings reveal that the preictal phase is marked by a gradual increase in instability, whereas the onset of the ictal phase is characterized by a sharp decrease in instability, followed by a substantial rise. The temporal evolution of cortical stability shows minimal within-subject variability, in contrast to the significant within-subject variability observed in cortical chaos. Furthermore, our results indicate that inter-regional communication acts as the chaotic behaviour driver in the epileptogenic zones during focal epileptic seizures.

## 2. Methods

### 2.1. Dataset

This study analyzed focal epileptic seizures in the NeuroVista dataset [35], which includes long-term iEEG data from 15 subjects. The patient cohort and the methodology for collecting their chronic iEEG data have been comprehensively detailed in the prior publication [35]. To summarize, all participants were diagnosed with refractory focal epilepsy, experiencing 2–12 seizures monthly. For data acquisition, each subject underwent implantation with 16 surface iEEG electrodes strategically positioned over the brain quadrant identified as the probable epileptogenic zone. The data used in this paper was approved by Human Research Ethics Committee, St Vincent’s Hospital Melbourne, approval LRR145/13.

Seizures from 12 NeuroVista subjects were examined to assess seizure pathways and cortical dynamics across extended timescales in individuals with focal epilepsy. Subjects 5, 12, and 14 were excluded due to an insufficient number of seizures. Additional patient details are provided in Appendix B1.

### 2.2. Single population model

To derive a population model, we begin by defining the mean membrane potential of a neural population, *v*_*n*_, as the sum of contributing mean post-synaptic potentials, *v*_*mn*_, where the post-synaptic and pre-synaptic neural populations are indexed by *n* and *m*, respectively. Each post-synaptic potential arises from the convolution of the input firing rate, *ϕ*_*m*_(*t*), with the post-synaptic response kernel,

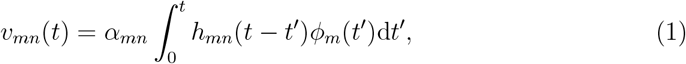

where *α*_*mn*_ is a lumped connectivity parameter that incorporates the average synaptic gain, the number of connections and the average maximum firing rate of the presynaptic populations. All lumped connectivity parameters are assumed to be unknown, so must be inferred from data. The post-synaptic response kernels, denoted by *h*_*mn*_(*t*), describe the profile of the post-synaptic membrane potential of population *n* that is induced by an infinitesimally short pulse from the inputs (like an action potential). The post-synaptic response kernels are parameterized by the time constant *τ*_*mn*_ and are given by

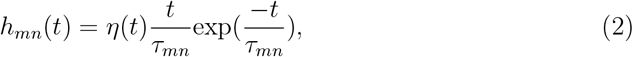

where *η*(*t*) is the Heaviside step function. Typically, *α*_*mn*_ and *τ*_*mn*_ are assumed to be constants (particularly for current-based synapses) that define the presynaptic population type. For the model that we are considering, the index *m* and *n* can represent either pyramidal (p), excitatory interneuron (spiny stellate) (e) or inhibitory interneuron (i) populations.

The inputs to the population, *ϕ*_*m*_, may come from external regions, *µ*, or from other populations within the model, *g*(*v*_*m*_), where

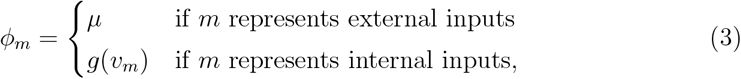

where internal inputs refer to the synaptic inputs that a population of neurons receives from other populations within the same model, while external inputs refer to synaptic inputs from regions outside the model. The various populations within the model are linked via the activation function *g*(·) that describes a mean firing rate as a function of the pre-synaptic population’s mean membrane potential. The activation function exploits a sigmoidal relationship between the mean membrane potential and firing rate of each of the populations. This sigmoidal nonlinearity may take different forms, but for this study, the error function form is used [21],

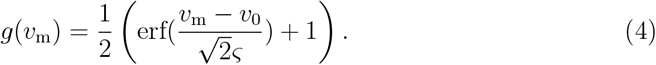

The quantity *ς* describes the slope of the sigmoid or, equivalently, the variance of firing thresholds of the presynaptic population (assuming a Gaussian distribution of firing thresholds). The mean firing threshold relative to the mean resting membrane potential is denoted by *v*_0_. The parameters of the sigmoidal activation functions, *ς* and *v*_0_, are usually assumed to be constants.

The convolution in Eq (1) can be written as two coupled, first-order ordinary differential equations, which is a second-order state-space model [21]. This gives the system,

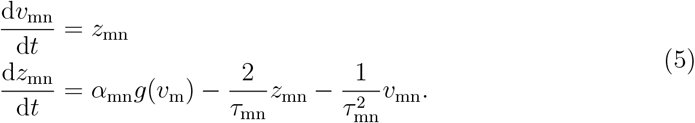

In summary, this single neural population model maps from a mean pre-synaptic firing rate to a post-synaptic potential. The terms that are usually considered parameters of the model are the synaptic time constants *τ*_*mn*_, the connectivity constants *α*_*mn*_, the mean firing thresholds *v*_0_, and firing threshold variances *ς*. These parameters can be set to describe connections between specific neural populations, such as pyramidal neurons, spiny stellate cells and fast and slow inhibitory interneurons.

### 2.3. Multiple populations for a single region model

Multiple populations in the form of Eq (5) can be configured and interconnected to represent the circuitry of a cortical region, such as a cortical column. Each synaptic connection in the model is described by the set of coupled first-order ODEs of Eq (5); however, the parameters are connection-specific. An illustration of the neural mass model used in this study is shown in Figure 1b. The formulation of the model is derived from the model introduced by Jansen and Rit [36], and has also been outlined in previous works [34, 37, 21, 38]. The neural mass model is suitable to model iEEG measured at this scale (electrodes approximately 5 mm in diameter with spacing on the order of cm), in line with similar neural models used to describe EEG/MEG activity [39, 29, 40].

**Figure 1.**
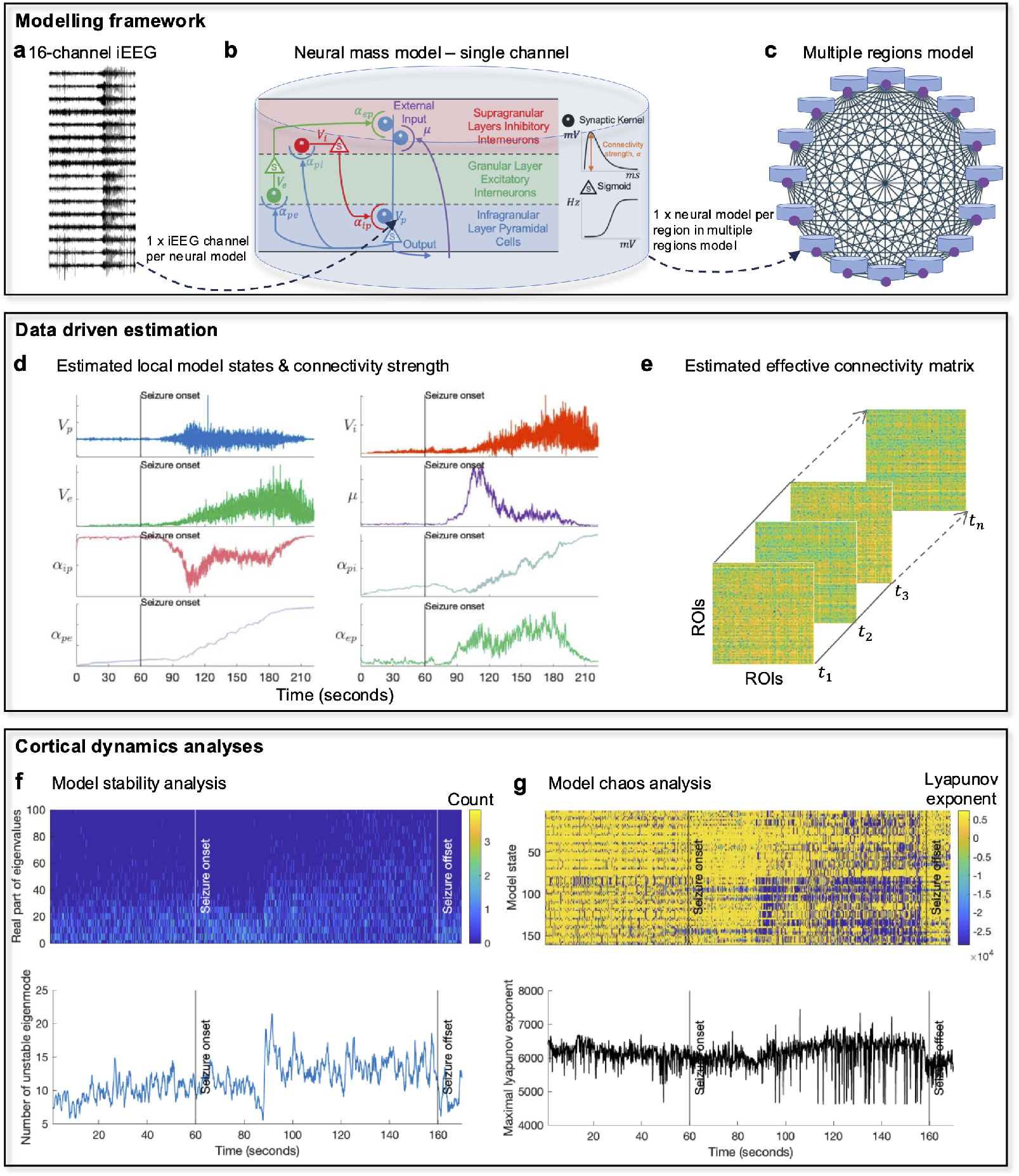
Conceptualization of the analysis framework. **a** Example of an iEEG recording of an epileptic seizure. **b** Schematic of the neural mass model. **c** Schematic of the multiple regions models consisting of 16 fully connected neural mass models. **d** Example estimation time series of local model states and connectivity parameters for one neural mass model in the multiple regions model. **e** Example estimation time series of effective connectivity strengths between local neural mass models. **f** Top: the time-varying eigenvalue spectrum of the multiple regions model’s Jacobian matrix for the example iEEG. Bottom: the time course of the number of unstable eigenmode. **g** Top: the time-varying Lyapunov spectrum of the multiple regions model for the example iEEG. Bottom: the time course of the maximal Lyapunov exponent.

The model comprises three neural populations, namely excitatory, inhibitory, and pyramidal populations (Figure 1b). The pyramidal population (in infragranular layers) driven by the external input *µ*, excites the spiny stellate excitatory population (in granular layer IV) and inhibitory interneurons (in supragranular layers) and is excited by the spiny stellate excitatory population and inhibited by the inhibitory interneurons. The model state vector is a concatenation of discrete time values of the post-synaptic membrane potentials and the derivatives of the potentials (see explanations of model variables in Table 1),

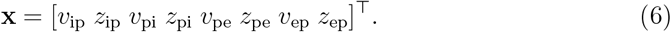

**Table 1.** List of variables used in the multiple regions model.

| Variable | Description |
| --- | --- |
| $v_i$ | Membrane potential of inhibitory neuronal population in neural mass model |
| $v_e$ | Membrane potential of excitatory neuronal population in neural mass model |
| $v_p$ | Membrane potential of pyramidal neuronal population in neural mass model |
| $\mu$ | Post-synaptic membrane potential induced by firings from other regions |
| $u$ | Time derivative of $\mu$ |
| $v_{ip}$ | Post-synaptic membrane potential from inhibitory to pyramidal population |
| $z_{ip}$ | Time derivative of $v_{ip}$ |
| $v_{ep}$ | Post-synaptic membrane potential from excitatory to pyramidal population |
| $z_{ep}$ | Time derivative of $v_{ep}$ |
| $v_{pi}$ | Post-synaptic membrane potential from pyramidal to inhibitory population |
| $z_{pi}$ | Time derivative of $v_{pi}$ |
| $v_{pe}$ | Post-synaptic membrane potential from pyramidal to excitatory population |
| $z_{pe}$ | Time derivative of $v_{pe}$ |
| $z_{pe}$ | Time derivative of $v_{pe}$ |
| $z_{pe}$ | Time derivative of $v_{pe}$ |
| $z_{pe}$ | Time derivative of $v_{pe}$ |
| $\alpha_{ip}$ | connectivity strength from inhibitory to pyramidal population |
| $\alpha_{pi}$ | connectivity strength from pyramidal to inhibitory population |
| $\alpha_{pe}$ | connectivity strength from pyramidal to excitatory population |
| $\alpha_{ep}$ | connectivity strength from excitatory to pyramidal population |
| $W$ | inter-regional connectivity matrix |
| $W_{ab}$ | inter-regional connectivity strength from region a to b |

The model parameter vector contains the external input and all connectivity strengths,

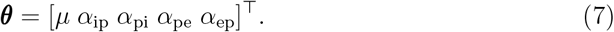

The dynamics for the parameter are modeled as a random walk,

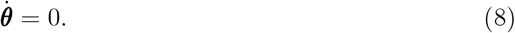

The state vector **x** and the parameter vector *θ* are concatenated to form the augmented state vector,

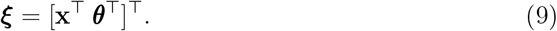

The augmented state space model is given by

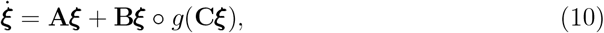

where ° denotes element-wise multiplication. The matrices **A, B**, and **C** are defined in Appendix A.1.

It is necessary to discretize the model for estimation purposes. The Euler method was used for discretizing the model. For the Bayesian inference scheme, it is also necessary to model uncertainty in our model by an additive noise term. With the inclusion of the additive noise term **w**_*t*_ the discrete time augmented state space model is denoted by

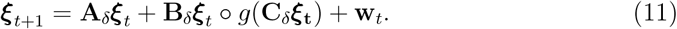

The model uncertainty is defined by a zero mean, temporally white Gaussian with known covariance matrix **Q**. In forward models, **w**_*t*_ is used as a driving term to simulate unknown input to the system from afferent connections or from other cortical regions. However, for model inversion purposes, this additional term also facilitates estimation and tracking of parameters via Kalman filtering or other Bayesian inference schemes. For the Kalman filter, the covariance of **w**_*t*_ quantifies the error in the predictions through the model. If we believe our model is accurate, then we set all of the elements of **Q** to a small value. On the other hand, a high degree of model-to-brain mismatch can be quantified by setting the elements of **Q** to larger values.

It is well accepted that the field potentials that are measured with iEEG are predominately generated by synaptic currents arising from inputs to the pyramidal neurons [41]. In our model, these currents are linearly proportional to the mean membrane potential of the pyramidal population. Therefore, the measurement is modeled as the mean membrane potential of the pyramidal population, which is the sum of the incoming post-synaptic membrane potentials. The measurement model is of the form,

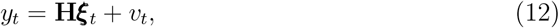

where 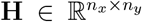 is the observation matrix, *v*_*t*_ ∼ 𝒩 (0, **R**). As our measurement function is linear, **H** is simply an index vector of zeros and ones that defines the average pyramidal membrane potential given by

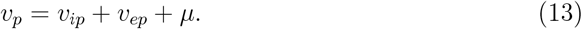

### 2.4. Semi-analytical Kalman filter

The aim of the Kalman filter is to estimate the most likely sequences of states 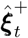 and the corresponding error covariances 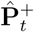,given knowledge of the biophysics and anatomy of the brain regions of interest combined with the noisy measurements *y*,

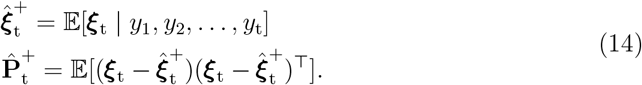

The Kalman filter proceeds in two steps: prediction and update. In prediction, the prior distribution is propagated through the neural mass model. This step provides the so-called *a priori* state estimate distribution,

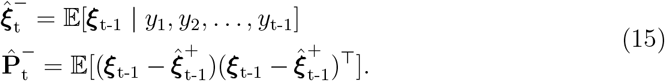

In the second step, a Bayesian update is performed to correct the *a priori* state estimate based on the actual measurement, giving the posterior distribution,

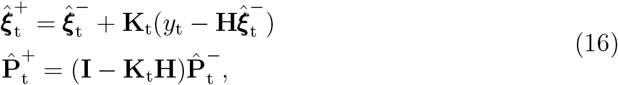

where **K**_t_ is the Kalman gain [42], a weighting to correct the *a priori* state estimate. The Kalman gain is calculated using the available information about the confidence in the prediction of the augmented states through the model and the observation model that includes noise by

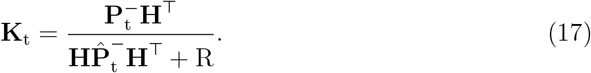

After each time step, the *a posteriori* estimate becomes the prior distribution for the next time step and the filter proceeds. See previous papers [34, 21, 38] for details about how to handle the nonlinearity of the model with the Kalman filter.

The Kalman filter requires 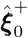 and 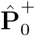 to be initialized to provide the *a posteriori* state estimate and state estimate covariance at time *t* = 0. The other parameters that must be initialized are the model and measurement noise **Q** and *R*, respectively. Further details of filter initialization are given in Appendix A.2.

### 2.5. Multiple regions model

Coupling of cortical region *b* to region *a* is achieved by connecting the output firing rate of the pyramidal population in region *b* to the input of the pyramidal population in region *a* via a post-synaptic response kernel of the same form in Eq (1). The inputs from the firing rates are modeled for every pyramidal population using the same form of second-order model defined in Eq (5). All interconnections between regions were assumed to have the same kernel, which was parameterized by a time constant, *τ*_*d*_ [43].

The firing rates form standard inputs to the pyramidal cells in all other cortical regions and induce post-synaptic potentials via a convolution kernel as described by Eq (1).

In the multiple regions model, each region is modeled by a neural mass model and the state vector has two extra states for the post-synaptic membrane potentials induced by firing rates from other regions. The state vector for region *a* is given by

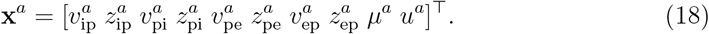

The dynamics for region *a* are described by

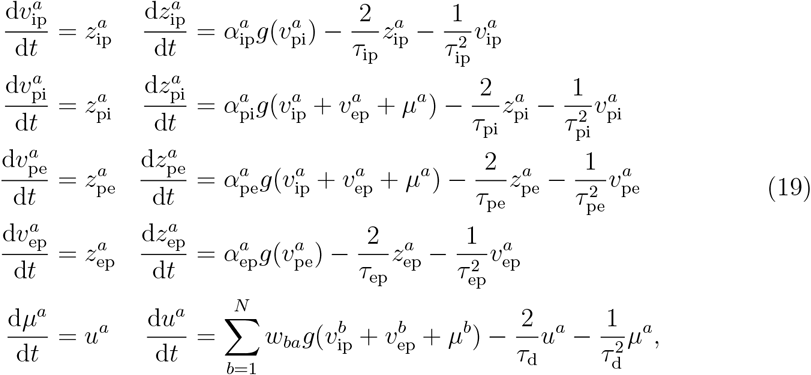

where the superscript *b* indexes the model in region *b*, and *w*_*ba*_ denotes the inter-regional connectivity strength from *b* to *a*. The state variable *µ* represents the post-synaptic membrane potential induced by inter-regional firing rates, and *u* is the time derivative of *µ*. Let **X** include all state variables of *n*_*y*_ regions 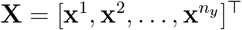,the multiple regions model is of the differential equation,

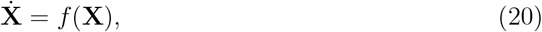

where *f* is a map from 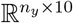 to 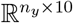 based on Eq (19).

The model parameter vector contains inter-population connectivity **Θ** for every region,

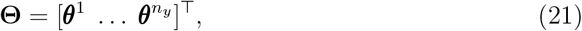

and inter-regional connectivity **W**,

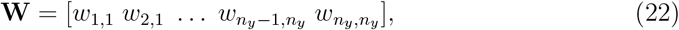

and they form a parameter vector,

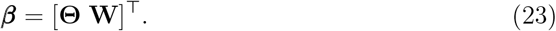

The augmented state vector for the model is of the form,

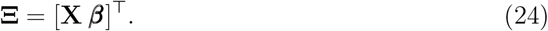

The augmented state space model is given by

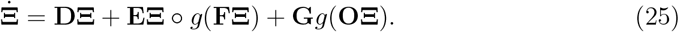

The matrices **D, E, F, G** and **O** are defined in Appendix A.3.

Similar to the single region model, the Euler method was used for discretizing the model. A noise **w** ∼ 𝒩 (0, **Q**) was added to the discrete time version of the model and is given by

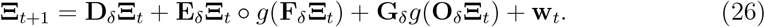

The measurement model is given as

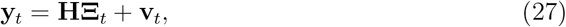

where 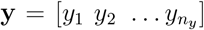 consists of measurements of *n*_*y*_ channels, 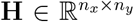 is the observation matrix, and **v**_*t*_ ∼ 𝒩 (0, **R**) is a zero mean, spatially and temporally white Gaussian noise. The matrix **H** defines a summation of the relevant membrane potentials (corresponding to excitatory and inhibitory postsynaptic responses of pyramidal populations) that contribute to each iEEG channel.

### 2.6. Multivariate linear regression

To estimate inter-regional connectivity strengths of the multiple regions model, we first treat each region in the model independently and apply the Kalman filter described in Section 2.4. The Kalman filter takes a single channel iEEG and estimates the sequences of the *a posteriori* augmented state distribution (including both model states and parameters) for the corresponding single region model. The external input *µ* in the single region model represents the summation of the post-synaptic membrane potential induced by inter-regional firing rates and is given by

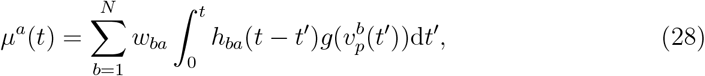

where the post-synaptic kernel *h*_*ba*_ is defined by Eq (2) with time constant *τ*_*d*_ and 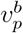 is the pyramidal membrane potential in region *b*. The inter-regional connection strength *w*_*ba*_ is estimated by the multivariate linear regression with the estimates of *µ* for all regions and the convolution between the kernel and the firing rates induced by the estimates of *v*_*p*_ for all regions. The reason we do not use Kalman filters to estimate the inter-regional connectivity is to keep the method computationally feasible.

The multivariate regression model relates more than one predictor and more than one response. Let **Z** be an *n* × *N* matrix, where each column is a *µ* estimate time series, and let **F** be an *n* × *N* matrix, where each column is a time series of the convolution of the post-synaptic kernel with *v*_*p*_ estimates. Let **B** be an *N* × *N* matrix of coefficients *w*_*ba*_ and let **E** be an *n* × *N* Gaussian white noise. The multivariate regression model is defined by

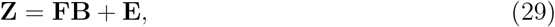

where the maximum likelihood estimation and unbiased estimator for **B** is given as

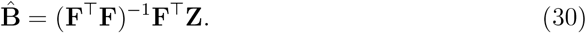

### 2.7. Local stability near the fixed points for the multiple regions model

We determine the fixed points, **X**^*^, by setting Eq (20) to zero 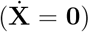 and solving for **X**. To identify a fixed point closest to our real-time parameter estimates, we initialize the solver using our parameter estimates as a starting point. To study the behaviour of the model near the fixed points, we linearize the multiple regions model and calculate its Jacobian matrix at the fixed point **X**^*^. The model comprises *N* regions and each region corresponds to one neural mass model with 10 dimensions. Therefore, the Jacobian **J** ∈ ℝ^10*N* ×10*N*^ is given by

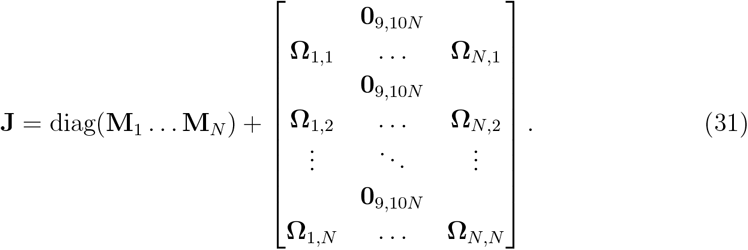

where 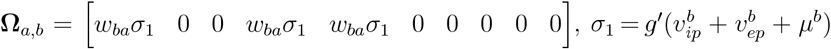,and **0**_*k,l*_ denotes the *k*-by-*l* all-zero matrix. diag(**M**_1_, …, **M**_*N*_) denotes the diagonal matrix with block components **M**_*a*_ ∈ ℝ_10×10_ defined as

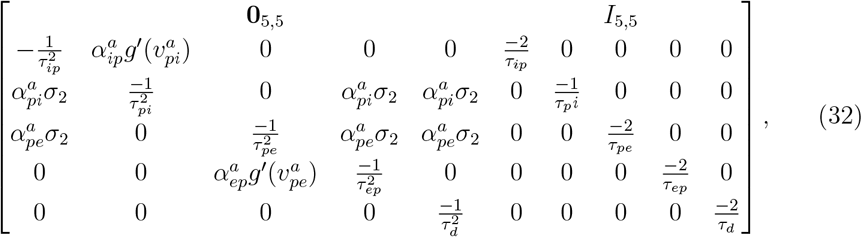

where 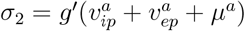.

The Jacobian is decided by the inter-population connectivity *α*_*mn*_ and the inter-regional connectivity *w*_*ba*_. We calculate the eigenvalues *λ*_*k*_ of **J** at the equilibrium point **X**^*^ to see if the system is stable around **X**^*^. Formally, we perform the eigenvalue decomposition **J** = *U* Λ*U* ^⊤^, where *U* is orthonormal, Λ is the diagonal eigenvalue matrix, and ^⊤^ means the transpose conjugate. We call the real part of *λ*_*k*_ the criticality index. For a linearized system, if the criticality index is less than 0, the corresponding mode is stable; a small perturbation along the eigenvector will decay and the system will return to the equilibrium point. Conversely, if the criticality index is greater than 0, even a trivial perturbation along the eigenvector will diverge the system from the equilibrium point. When the criticality index equals 0, the system is at the neutral point along the corresponding eigenvector. In short, the system is unstable if there is at least one unstable eigenmode, and is stable when all eigenmodes are stable.

The evaluation of the number of unstable eigenmodes is also crucial, as a higher count signifies increased instability, implying multiple independent directions for perturbation growth. The eigenvalues associated with unstable eigenmodes also reflect the degree of instability, as the real part of eigenvalues represents the growth or decay rate of perturbations.

In summary, linear stability refers to the tendency of a system to return to its equilibrium or steady state after a small perturbation, and it is often analyzed using linear stability analysis. Studying cortical stability in epilepsy is crucial for predicting seizure onset and understanding how small perturbations in brain activity can lead to large-scale disruptions.

### 2.8. Lyapunov spectrum of multiple regions model

Lyapunov exponents measure how two trajectories that start infinitesimally close in the phase space diverge or converge as time progresses [44]. If they diverge exponentially over time, it indicates the presence of chaos in the system. The Lyapunov exponent gives a quantitative measure of this rate of divergence or convergence.

To determine the Lyapunov exponent for the multiple regions model defined in Eq (20), consider a perturbation *δ***X**(*t*) to the trajectory **X**(*t*). The rate of change of this perturbation is governed by the Jacobian **J**(**X**(*t*)) of the vector field evaluated at the trajectory,

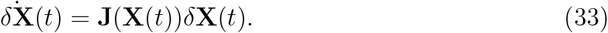

In a multidimensional system, the Lyapunov exponents are represented as a vector, ***λ*** = (*λ*_1_, *λ*_2_, …, *λ*_*n*_), where each *λ*_*i*_ characterizes the exponential rate of divergence or convergence along a particular direction in phase space. The maximal Lyapunov exponent, denoted as *L*_max_, is often used as a measure of the system’s predictability; it quantifies the largest average exponential rate of growth of a perturbation over time and is mathematically defined as

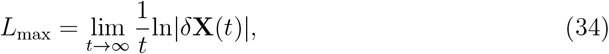

given an initial perturbation *δ***X**(0).

Specifically, for a *d*-dimensional phase space, there are *d* Lyapunov exponents. These are ordered as *λ*_1_ ≥ *λ*_2_ ≥ · · · ≥ *λ*_*d*_, and their collective set constitutes the spectrum of Lyapunov exponents. The spectrum depends on the starting point **X**(0). In our implementation, we calculated the Lyapunov spectrum at the equilibrium point, which is close to the model state estimate, and 1000 samples drawn from an isotropic Gaussian around the equilibrium point. An exponent with a positive value (*λ*_*i*_ *>* 0) indicates trajectories in the corresponding direction in phase space are diverging exponentially with time. An exponent valued at zero (*λ*_*i*_ = 0) points to neutral stability in that direction. A negative valued exponent (*λ*_*i*_ *<* 0) suggests trajectories in its corresponding direction are converging. Particularly, the largest Lyapunov exponent is of utmost importance. A positive value for this exponent implies the system’s sensitive dependence on initial conditions, a defining trait of chaos. In most cases, the computation of Lyapunov exponents cannot be carried out analytically so numerical calculation is performed.

In short, studying cortical chaos in epilepsy is important for understanding the complex, unpredictable dynamics of seizure activity and how small changes in brain state can lead to significant, erratic disruptions.

## 3. Results

We analyzed a total of 3008 seizures, recorded using 16-channel iEEG, from 12 patients with focal epilepsy who underwent chronic recordings as part of the NeuroVista seizure prediction study (average 251 seizures/patient) [35]. The dataset is available at https://www.epilepsyecosystem.org/neurovista-trial-1 [45]. Each seizure recordings started 1 min before seizure onset and ended 10 s after seizure offset. Figure 1a shows an example iEEG recordings of a focal epileptic seizure.

### 3.1. Conceptualization of analysis framework

We created a multiple regions model comprising 16 neural mass models (NMMs) fully connected to each other, as illustrated in Figure 1c. The mean (spatial not temporal) membrane potential of the pyramidal population in each NMM is considered proportional to the voltage of one iEEG channel. As shown in Figure 1b, the NMM consists of three neuronal populations within a region—excitatory pyramidal neurons, excitatory spiny stellate neurons, and inhibitory interneurons—interacting locally and across regions through long-range inter-regional connections. We estimated the temporal evolution of regional NMM neurophysiological variables for each region (Figure 1d) and long-range inter-regional synaptic connection strengths based on iEEG signals (Figure 1e).

We describe the dynamics of epileptic seizures from two perspectives. First, the time-evolved eigenvalue spectrum of the model’s Jacobian matrix around the fixed point (Figure 1f) provides insights into the linear stability of the system in each one-second time window, with eigenvalues indicating how perturbations from the fixed point evolve over time. To compute the eigenvalue spectrum of the model, we identified the fixed point given the parameter estimates in each time window, linearized the model around the fixed point, and computed the eigenvalues of the Jacobian matrix. Stable fixed points have eigenvalues with negative real parts only, while unstable points have positive real parts[46].

Second, the time-evolved Lyapunov spectrum of the model (Figure 1g) provides information about the system’s chaos over time. The Lyapunov spectrum of a dynamical system constitutes a set of exponents, each corresponding to distinct directions within the system’s phase space. A high chaos system is highly sensitive to initial conditions, leading to unpredictable and complex behavior, while a low chaos system is more stable and predictable [47].

The analysis framework employed herein is an inference-based time-resolved analysis of cortical dynamics based on multiple region models. Notably, our methodology involves semi-analytical calculation of cortical dynamics, specifically the model Jacobian eigenvalue spectrum and Lyapunov spectrum-using model parameter estimates derived from real clinical data. Generally, this analysis framework presents two principal advantages:

i. The assessment of the time-evolving spectra of Jacobian eigenvalues and Lyapunov exponents facilitates the identification of temporal variations in cortical stability and chaos during seizures. These identified variations hold the potential to serve as biomarkers if collective analyses yield statistically significant outcomes.
ii. The Lyapunov spectrum can pinpoint the chaotic drivers, denoting states consistently associated with the largest Lyapunov exponent [47]. In the context of epilepsy models, a chaotic driver signifies specific model states consistently contributing to chaotic dynamics, playing a pivotal role in generating intricate and unpredictable patterns inherent in epileptic seizures. Identifying chaotic drivers is imperative for elucidating and potentially intervening in the chaotic nature of epileptic occurrences.

### 3.2. Seizure trajectories

Figure 2 presents the parameter estimation results for the five local connectivity strengths and long-range effective connectivity. Overall, seizures across multiple events for each individual followed a relatively consistent trajectory through the parameter space of the multiple regions model, suggesting that seizure transitions adhere to a stereotypical pathway. Importantly, these transitions are synchronized with the time of seizure onset.

**Figure 2.**
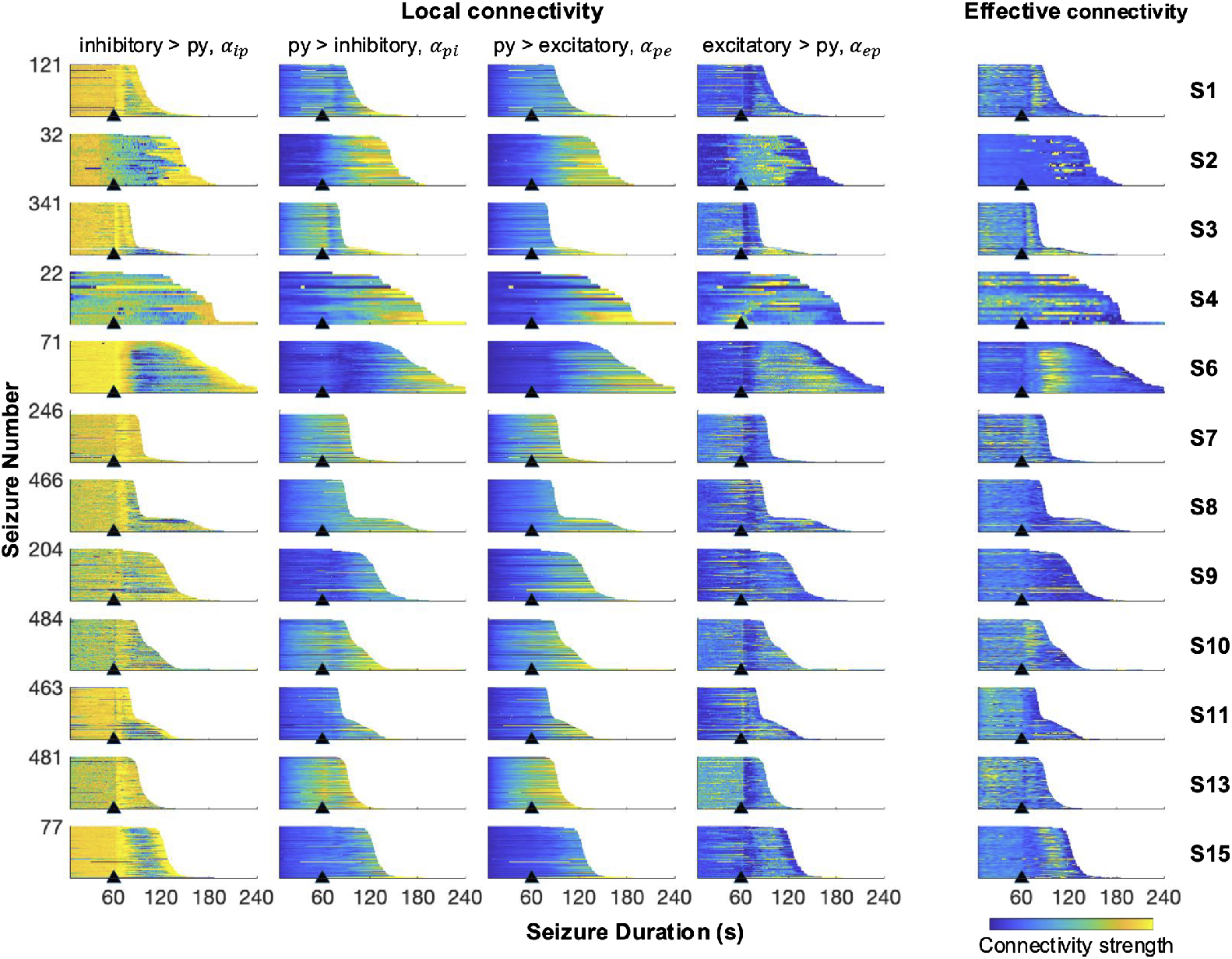
Estimated changes in parameter trajectories during every seizure. The figure illustrates the estimated local and effective connectivity parameters derived from seizure recordings across 12 subjects. Each row represents data from one subject, and the subpanels within each row display the parameter estimates across seizures, with the specific number of seizures indicated by the numbers on the left. The onset of each seizure is marked by a black triangle, and the data extends up to 10 seconds after the seizure offset. Averaged estimates across 16 channels are shown for local connectivity parameters, and averaged strength is shown for effective connectivity.

Across all patients, the most pronounced ictal changes in connectivity strength were observed in the in-going connections to the pyramidal population (i.e., *α*_*ip*_, *α*_*ep*_), as well as in the effective connectivity. Specifically, *α*_*ip*_ displayed a significant reduction following a brief increase shortly after seizure onset for most subjects. Effective connectivity, on the other hand, exhibited a marked increase post-onset in certain subjects, including S1, S6, S10, and S15. In contrast, the outgoing connections from the pyramidal population (i.e., *α*_*pi*_, *α*_*pe*_) remained relatively stable throughout the seizures. However, gradual and substantial increases were observed post-onset in subjects S2, S4, S6, and S13, indicating a delayed but notable change in these connections.

The subsequent sections leverage these parameter estimates to perform stability and chaos analyses.

### 3.3. Cortical instability first decreases then increases during seizures

Figure 3a illustrates the results of the stability analysis performed on the multiple regions model using an example iEEG data from subject 2. The top panel illustrates how the distribution of the Jacobian eigenvalues evolves, with a pronounced increase in density of unstable eigenvalues (indicated by warmer colors) coinciding with the seizure onset. This shift reflects a significant destabilization of the system. As the seizure progressed, the density of unstable eigenvalues fluctuated but generally remained elevated until the seizure offset, at which point the distribution shifted back toward greater stability. The bottom panel quantifies this instability, showing a marked rise in the number of unstable eigenmodes at seizure onset, indicating the system’s transition into a hyperexcitable state. This heightened instability persisted throughout the ictal phase and diminished rapidly upon seizure termination, aligning with the restoration of neural homeostasis. These findings underscore the utility of stability analysis in capturing the nonlinear dynamics of seizure evolution and highlight how sudden transitions to instability may underpin the emergence and termination of epileptic events.

**Figure 3.**
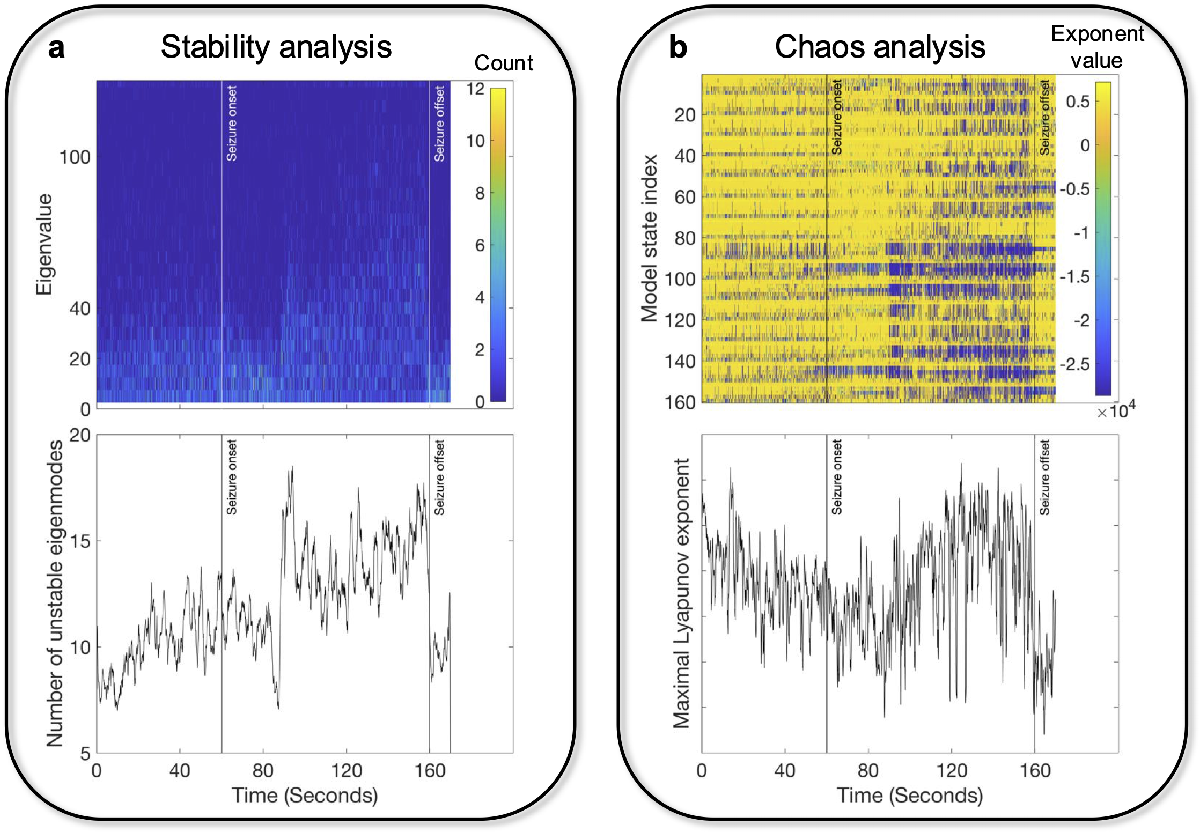
Stability and chaos analysis with an example iEEG recording. **a** Top: evolution of the distribution of the Jacobian’s eigenvalues as a function of time from 60 seconds before seizure onset to 10 seconds after seizure offset. Yellow denotes high density, while blue denotes low density. Bottom: The corresponding number of unstable eigenmodes as a function of time. **b** Top: evolution of Lyapunov spectrum as a function of time from 60 seconds before seizure onset to 10 seconds after seizure offset. For each time window, the magnitude of exponents are shown by color. Yellow means large exponent, while blue means low exponent. Bottom: The corresponding maximal Lyapunov exponent in each time window.

We extended our analysis to a group level by examining 12 subjects with multiple recordings, using the same methods to uncover shared or unique stability patterns. Figure 4a shows the temporal evolution of the number of unstable eigenmodes during each seizure event for each subject. The temporal variation pattern of dynamic stability, in terms of unstable eigenmodes, was remarkably consistent across events for each subject, suggesting that within-subject cortical stability transitions follow a stereotypical manner.

**Figure 4.**
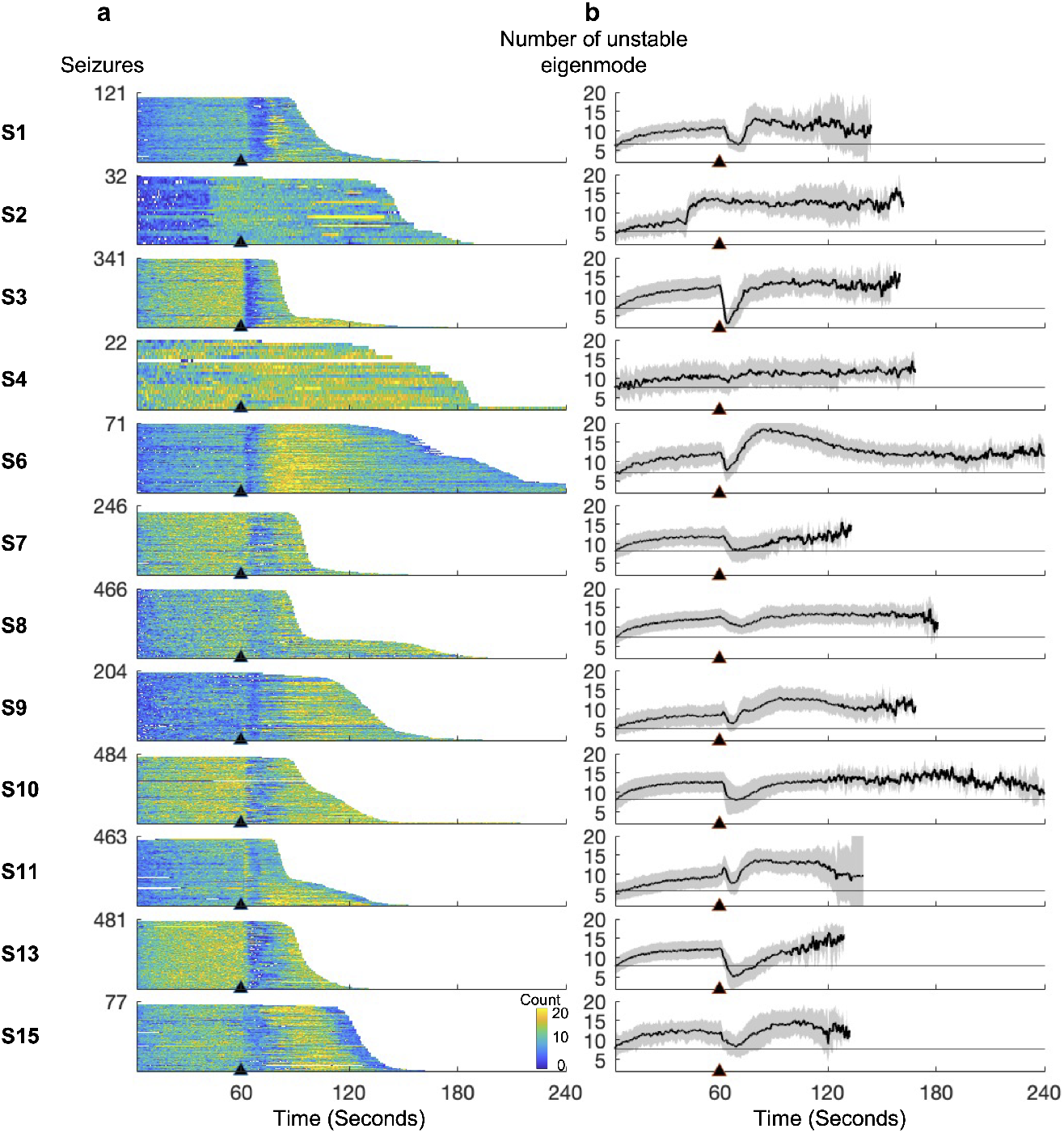
Changes in the number of unstable eigenmodes during epileptic seizures. **a** Each subpanel represents the time-varying number of unstable eigenmodes in the multiple regions model’s Jacobian matrix spanning all seizures for each subject. Seizures are sorted shortest to longest. **b** Each subpanel represents the mean number of unstable eigenmodes (averaged across seizures) for each subject (grey shading is the 95% confidence interval of the mean). The horizontal gray line represents the baseline (the number of unstable eigenmodes at the beginning of the recording).

Figure 4b presents the mean change in the number of unstable eigenmodes during seizures. For all subjects, the average number of unstable eigenmodes increased from the baseline (value at the beginning) in the preictal period, indicating that cortical dynamics instability consistently rises before seizure onset, causing the system to deviate from equilibrium more rapidly with minor perturbations. In all subjects except S2 and S4, there was a substantial drop in the number of unstable eigenmodes immediately after seizure onset, with some subjects showing numbers below the baseline. This suggests that the onset of a seizure represents a spatiotemporal transition from a very unstable state to a less unstable state. Subsequently, the degree of instability increased immediately after the decline and was highest during the ictal phase. This was confirmed in all subjects except S13 by paired t-tests (p *<* 0.01), which showed that the stability measure in the ictal period was significantly greater than in the preictal period. Overall, cortical activity in epileptogenic regions exhibited higher instability during seizures compared to pre-seizure periods, despite a significant transient decrease at the beginning of seizures.

### 3.4. Seizures as individualized changes in maximal Lyapunov exponent

Figure 3b presents the temporal evolution of the Lyapunov spectrum for the model based on an example iEEG recording. The top panel shows the Lyapunov spectrum and a substantial decrease in the magnitude of the Lyapunov exponents (indicated by the transition from yellow to blue) occurred around seizure onset, signifying an reduction in the system’s chaotic dynamics. This decreased chaos persisted throughout the early phases of the seizure. A noticeable growth of chaos was observed subsequently denoted by the transition from blue to yellow, meaning the increased chaotic dynamics over the epileptogenic regions. The bottom panel depicted the temporal evolution of the maximal Lyapunov exponent, *L*_max_, during the seizure. It consistently decreased from a minute before the seizure onset to around 10 seconds after seizure onset. The exponent then observed a sharp increase to a high level and maintained that during the rest of the seizure, ending with a notable drop at the seizure offset.

We expanded our analysis to include 12 subjects, applying the same chaos metrics to identify consistent patterns or variations across the group. Figure 5a illustrates the within-subject variability in the temporal evolution of *L*_max_ for all subjects. Seizures in subjects S1, S3, S6, S7, S11, S13, and S15 followed a consistent evolution of *L*_max_, indicating that the chaos of cortical activity during seizures transitions in a stereotypical pathway. Conversely, subjects S2 and S4 showed noticeable within-subject variability in the temporal change of *L*_max_, while subjects S8, S9, and S10 exhibited negligible changes in chaos during seizures.

**Figure 5.**
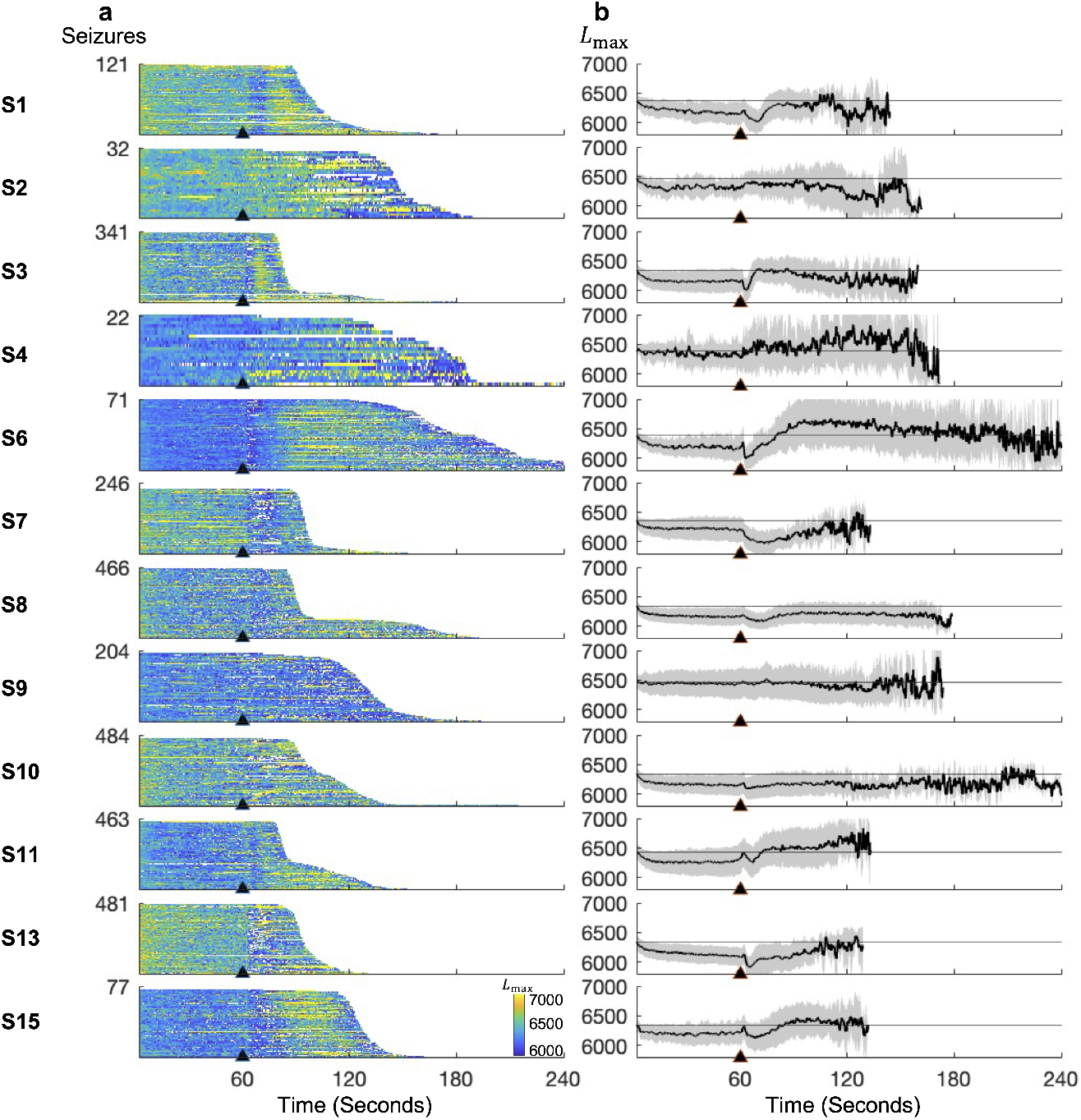
Change in maximum Lyapunov index during each epileptic seizure. **a** Each subpanel represents the time-varying *L*_max_ of the multiple regions model for all seizures for each subject. Seizures are sorted from shortest to longest. **b** Each subpanel represents the mean *L*_max_ (averaged across seizures) for each subject (grey shading is the 95% confidence interval of the mean). The horizontal gray line represents the baseline (*L*_max_ at the beginning of the recording).

Figure 5b shows the mean change of *L*_max_ during seizures across all events for each subject. Some general trends can be identified in most subjects, although no universal pattern was found in the mean changes of *L*_max_ across all subjects. A consistent motif was that *L*_max_ continuously decreased during the preictal phase for most subjects except for S4 and S9, who showed negligible changes in *L*_max_. This suggests that the preictal state represents a spatiotemporal transition from a more chaotic to a less chaotic state. Following seizure onset, there was a more noticeable decline in *L*_max_ for most subjects except S2, S4, and S9. The more ordered cortical activity likely reflects the synchronized rhythmic firing patterns of neurons involved in epileptic discharges. The remaining ictal period saw a reset of cortical dynamics from ordered to chaotic, evidenced by the increasing *L*_max_ in most subjects except S9. While most subjects showed *L*_max_ values exceeding the preictal baseline, the *L*_max_ of S8 remained significantly below the baseline.

### 3.5. Exogenous input driving epileptic dynamics between chaos and order

We computed the temporal evolution of the Lyapunov spectrum of the multiple regions model for each seizure (Figure 6a). This approach facilitated the identification of model states associated with *L*_max_ during seizures. For this, we determined the model state index corresponding to *L*_max_ in each time window (Figure 6b). Since the multiple regions model consists of interconnected neural mass models that are identical in both architecture and types of model states, we identified the specific model state associated with *L*_max_ in each time window (Figure 6c). Figure 6d illustrates the frequency of each variable associated with *L*_max_ across the entire recording. In this example, state variable *z*_ep_, representing the time derivative of the post-synaptic membrane potential from the excitatory to pyramidal neuronal population, corresponded to *L*_max_ in most time windows. This indicates that *z*_ep_ was primarily responsible for the cortical chaotic behaviours and played a significant role in driving epileptogenic zones between order and chaos.

**Figure 6.**
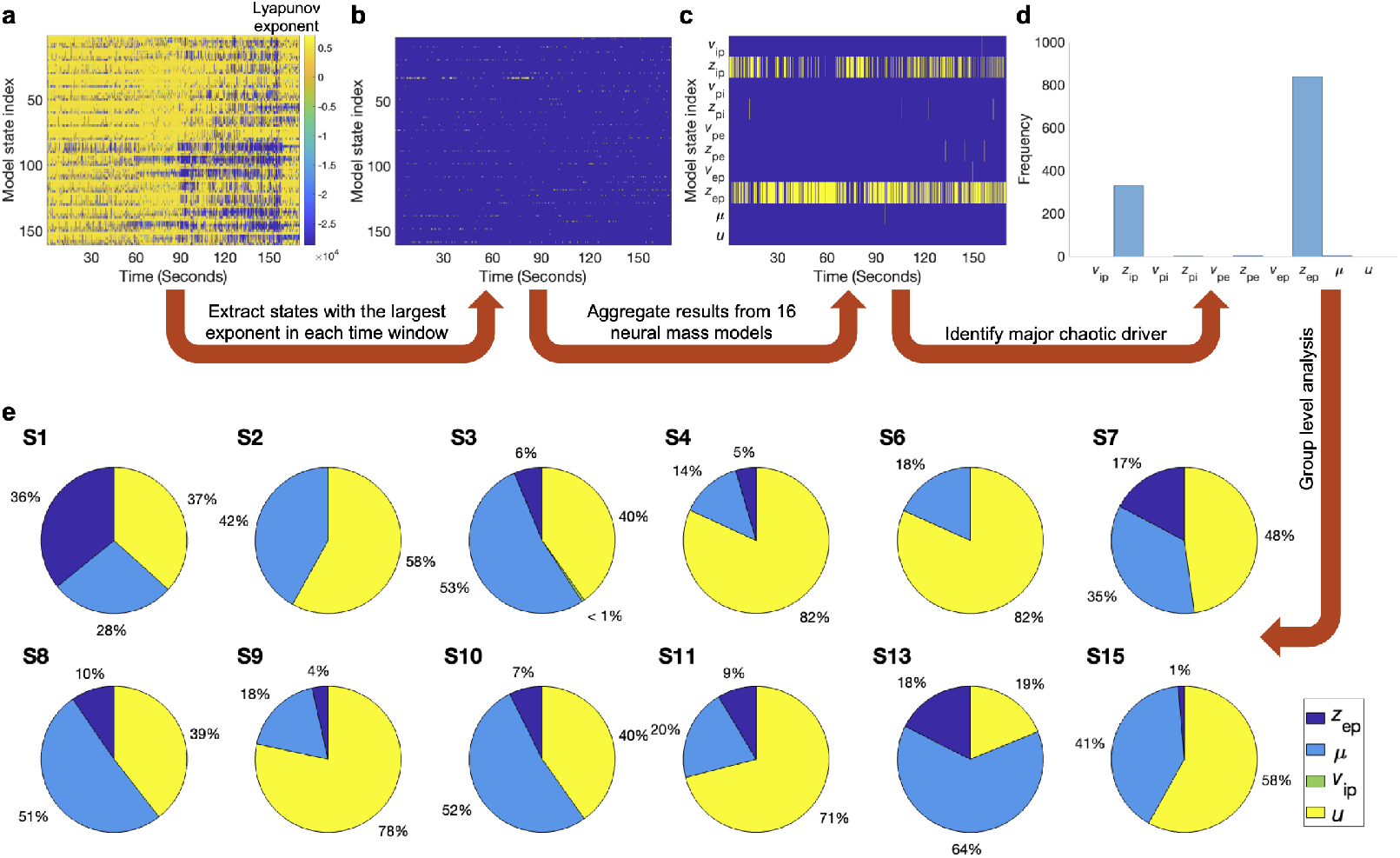
Identification of cortical chaotic driver. **a** Time-evolved Lyapunov spectrum. Abscissa indicates time and ordinate refers to multiple regions model states. Blue to yellow indicates small to large Lyapunov exponent values. **b** Highlighted points on each column indicate the chaotic driver corresponding to the maximal Lyapunov exponent in each time window. **c** Highlighted points on each columns are the type of model state associated with the maximal Lyapunov exponent in each time window. **d** The histogram shows the model state serving as the chaotic driver during the whole seizure recording. The dominant chaotic driver is the one with the highest frequency. **e** The percentage of model states that are the dominant chaotic driver in each subject is shown.

Group-level analysis in Figure 6e revealed that, among the 12 subjects, eight demonstrated the time derivative of the post-synaptic membrane potential induced by inter-regional neuronal firing, *u*, as the primary contributor to chaotic behavior in the epileptogenic regions. The remaining four subjects showed that the post-synaptic membrane potential resulting from neuronal firing between regions, *µ*, primarily influenced the chaotic dynamics observed. Notably, *µ* and *u*, as the primary drivers of chaos, were predominant in over 80% of seizures in all subjects except for S1, where *z*_ep_ accounted for one third of seizures. Nevertheless, *µ* and *u*, representing exogenous inputs from other regions, are the main neurophysiological processes in steering the degree of cortical chaos in focal epilepsy while the time-derivative of post-synaptic membrane potential from excitatory to pyramidal population *z*_*ep*_ also serves as chaotic driver in a noticeable number of seizures in 9 of 12 subjects.

## 4. Discussion

In this study, we analyzed temporal evolution of the stability and chaos of cortical dynamics in human epileptogenic brain region networks in a period from 60 seconds prior to seizure onset to 10 seconds after seizure offset in drug-resistant patients undergoing iEEG monitoring. We hypothesized that the degree of stability and chaos of cortical dynamics within the clinically identified epileptogenic regions would vary noticeably prior to, and post, seizure onset. We implemented and fit a multiple regions model consisting of 16 interconnected neural mass models to the data, and estimated model state and parameters over time. The model and parameter estimates allowed us to track the cortical stability and chaos of the epileptogenic brain regions.

For dynamic stability, the consistent pattern over time across seizure events for each subject, as shown in Figure 4a, underscores a stereotypical transition in cortical stability within individuals. This consistency highlights the potential for predicting and understanding the onset and progression of seizures in a subject-specific manner. Figure 4b illustrates an increase in the number of unstable eigenmodes in the preictal period across subjects. This verified an established theory of increasing instability of cortical dynamics preceding epileptic seizures [48, 49, 50, 51, 52]. The difference is that we directly calculated the Jacobian matrix eigenvalues of the multiple regions model using parameter estimates from the clinical data, enabling a fine-grained temporal resolution picture of the evolution of the dynamical stability spectrum to describe the epileptogenic regions. Whereas other studies mainly examined the stability from the response (e.g., line length) of models to active probings or spontaneous perturbations [53]. Furthermore, the progressive loss of model stability marks dynamic features of critical slowing in the transition to focal seizures [54, 55]. Critical slowing down in the context of seizure is that the brain becomes sensitive to perturbations in an invisible way because of very subtle changes in a slow underlying process [55, 54, 56]. Many neurophysiological processes have been identified and hypothesized to play crucial roles in epileptic seizure development [57, 21]. Nonetheless, most of these processes may simply represent random perturbations that trigger epileptic seizures in neural networks whose dynamics are already approaching catastrophic bifurcation through critical slowing down on certain timescales.

We also demonstrated that a noticeable reduction of instability at the start of seizure onset followed by a rise of instability. This observation can be supported by two aspects from human and animal experiments. First, excess synchronization of neuronal activity at the onset of a seizure [58, 59] can lead to a more stable, organized pattern of activity, at least in the short term. Second, an ictal discharge is preceded by exhausted presynaptic excitatory neurotransmitter release [60], which temporarily reduces overall excitability and potentially promotes a less unstable state. Moreover, the involvement of inhibition is dominant at the start of seizures and thus prevents seizure spread initially even though neurons experience a period of large rhythmic depolarizations [61, 62]. We observed up to tens of seconds of decreasing instability before it increased, which matched the experimental observation after focal injections of GABAergic blockers to create an ictal focus [61]. This indicates that the shift from the preictal to the ictal state in spontaneous complex partial seizures does not occur instantaneously but via multiple complicated neurophysiological processes that need to be explored to establish a more comprehensive mechanism of regulating the evolution of seizures. The subsequent increase of instability of cortical dynamics up until the end of seizures aligns with observations made in various epilepsy studies [63, 64, 65, 52]. As the seizure progresses, it may lead to increased synchronization, hyperexcitability, and a breakdown of the normal balance between neuronal inhibition and excitation, contributing to heightened instability [66, 67, 68]. Our analysis showed a fine-grained dynamic stability of focal seizures and unveiled a seldom-observed abrupt decline in instability at the initiation of seizures.

The observation of the within-subject consistency in the evolution of chaos aligns with studies that suggest individualized patterns in seizure dynamics. Previous studies indicated that, while seizure characteristics can vary widely among individuals, they often follow a consistent pattern within the same individual over time [69, 70, 71, 72]. An interesting finding is that epileptic seizures manifested in opposite ways, with some individuals exhibiting more chaotic and unpredictable patterns while others exhibited more stable, predictable patterns. This coincides with an important finding that the seizure dynamics might not exhibit a large change of chaos in certain scenarios, such as specific seizure types or under certain input conditions [73, 74].

The decrease in *L*_max_ during the preictal phase, indicating a transition to less chaotic states, is in line with studies that have identified changes in brain chaos preceding seizures [75, 25, 24]. The sharp decline in *L*_max_ following seizure onset in some subjects reflects a transition to more synchronized neural activity, which is a well-documented characteristic of the seizure onset from previous studies [25, 24, 76, 58]. The increase in *L*_max_ during the remaining ictal period, indicating a return to highly chaotic brain state, confirmed observations directly derived from EEG signals [25, 24]. This observation is attributed by increase in brain complexity [77] and changes of cortical input frequency [73].

We found the critical roles of the exogenous input from neighboring and distant regions (*µ*) and its time derivative (*u*) in driving chaotic behavior within the cerebral cortex of the epileptogenic zone in epileptic seizures. The predominance of *µ* and *u* in over 80% of seizures across the majority of subjects underscores their significant influence on cortical dynamics. There is hitherto no study specifically uncovering chaotic dynamic regulation through inter-regional cortical communication during seizures, although biomolecular studies found changing neuromodulatory inputs could relate to chaotic behavior in the cerebral cortex [78] and dynamic regulation of inter-regional cortical communication during working memory tasks which may contribute to chaotic neuronal activity in cerebral cortex [79]. Notably, the exception observed in subject S1, where the time derivative of the postsynaptic membrane potential from excitatory populations to pyramidal populations played a substantial role, suggests a variability in the dynamical drivers of epileptic seizures that warrants further investigation. While studies highlighted the importance of excitability in modulating seizure dynamics from a biomolecular perspective [12, 80], the importance of excitability in driving chaotic behaviour during epileptic seizures has not been observed before. These findings suggest that exogenous inputs and changes of excitability are pivotal in steering the degree of chaos in focal epilepsy.

In terms of the methodology, to address the Kalman filter initialization, we followed the approach described in a previous paper [21], using parameter values corresponding to non-seizure brain activity, since the recordings began in a non-seizure state. We also excluded the first five seconds (2000 time points at 400 Hz) from our analysis, ensuring the filter had converged during this initial period, as the parameter estimates experienced drastic changes during this initial period and then stabilized, suggesting the filter had converged. Regarding the verification of our inference method, forward simulations have been done using parameter estimates showing reasonably negligible prediction error (see Figure C1). Effective connectivity was estimated using linear regression, which provided a good fit to the data.

The scope of our study was confined to individuals with drug-resistant focal epilepsy, raising uncertainties about the generalizability of our findings on dynamic stability and chaos to other epilepsy variants. Additionally, the adequacy of our data set, particularly for patients with fewer than 100 recordings, remains a point of potential concern. A further constraint of our methodology pertained to the identification of seizure commencement and conclusion, which were determined either through clinical judgment or algorithmic processes. The clinical labeling of seizures is inherently subjective and prone to variability among clinicians. Lastly, we acknowledge the limitations of the current parameter estimation approach, where effective connectivity is derived based on local inference for each neural mass model. In future work, we aim to develop a unified inference framework that allows for the simultaneous estimation of both local parameters and effective connectivity.

Our findings on the temporal evolution of cortical stability and chaos in epileptogenic brain regions open up several applications. By identifying consistent patterns in stability and chaos dynamics within individuals, we can explore personalized therapeutic strategies, such as targeted electrical or optogenetic stimulation, to preemptively counteract the progression toward a seizure state. The analysis of the chaotic drivers, specifically the roles of exogenous inputs and changes in excitability suggests an application in the design of adaptive neuromodulation therapies. By identifying and targeting these key drivers, clinicians could develop responsive systems that dynamically adjust neuromodulatory inputs in real time to regulate cortical chaos and prevent seizures. For instance, closed-loop brain stimulation devices could be programmed to monitor chaotic behavior and deliver electrical or pharmacological interventions precisely when chaotic drivers surpass a critical threshold, thereby stabilizing brain activity and minimizing the risk of seizure onset. Additionally, our observations could pave the way for novel diagnostic tools that use the dynamic spectrum of stability and chaos to better characterize seizure types [81], providing a deeper understanding of individual seizure mechanisms and enhancing treatment approaches in a dynamical systems context.

## Data Availability

All data produced in the present study are available upon reasonable request to the authors
All data produced in the present work are contained in the manuscript
All data produced are available upon reasonable request to the authors

## Ethical Statement

iEEG data were continuously recorded from individuals with refractory focal epilepsy using the NeuroVista Seizure Advisory System, an implanted device previously described in detail [35] (approved by the Human Research Ethics Committee of St. Vincent’s Hospital, Melbourne, under protocol LRR145/13).

## Code and Data Availability

The MATLAB code of the multiple regions model, parameter estimation, and stability and chaos analysis are available on GitHub (https://github.com/yundumbledore/NeuroProcImager-Pro). The dataset is available at https://www.epilepsyecosystem.org/neurovista-trial-1.

## Acknowledgments

This work is supported by grants from the Australian Research Council (DP210100045, DP200102600), National Health and Medical Research Council (GNT1183119), and Monash University. We thank MASSIVE (https://www.massive.org.au) for computational resources.

## Appendix A. Details of model

### Appendix A.1. Definition of matrices A, B, and C in neural mass model

The matrix **A** has a block diagonal structure that is comprised of two sub-matrices,

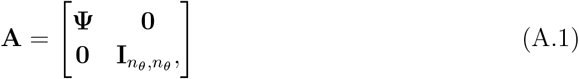

where 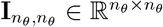 is the identity matrix and *n*_*θ*_ is the number of parameters in the model, and 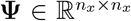 is also composed of the sub-matrices and *n*_*x*_ is the number of states in the model,

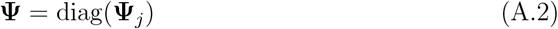

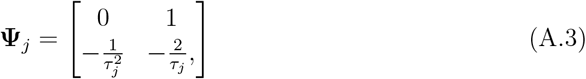

where *j* = 1, …, *N* indexes connections.

The discrete time version **A**_*δ*_ is related to **A** by

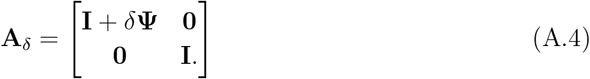

The matrix **B** has the form

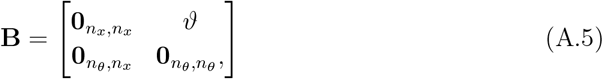

where 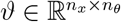 maps the connectivity to the relevant sigmoidal function and is of the form

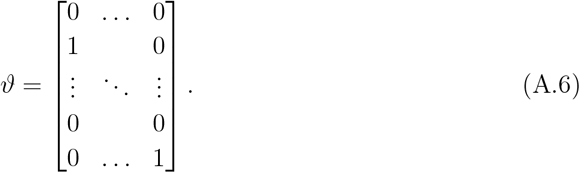

The discrete time version is simply

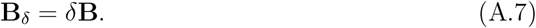

The adjacency matrix **C** is the same for both the continuous and discrete version of the model. It has a block diagonal structure where

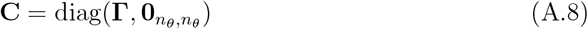

and 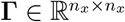 sums the relevant post-synaptic potentials to form the mean membrane potentials then maps them to the sigmoidal function and is of the form

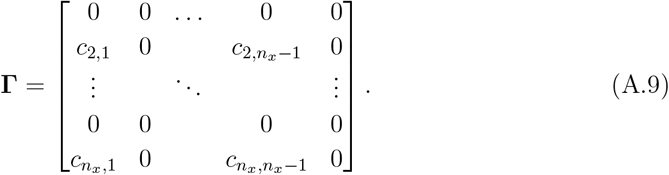

### Appendix A.2. Filter initialization

We used the average and covariance of simulated data to initialize 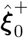 and 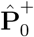. For the model uncertainty we used small constant values (5*µV*), which prevents the filter converging, and enables new measurements to continue to influence the estimation. For the measurement noise we used a value of 1*µV*.

### Appendix A.3. Definition of matrices D, E, F, G and O in the multiple regions model

The matrix **D** has a block diagonal structure that is comprised of two sub-matrices,

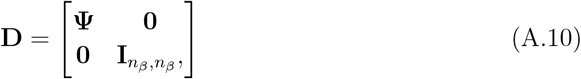

where 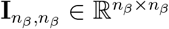 is the identity matrix and *n*_*β*_ is the number of parameters in the model, and 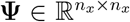 is also composed of the sub-matrices and *n*_*x*_ is the number of states in the model,

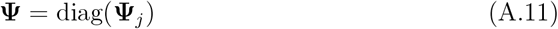

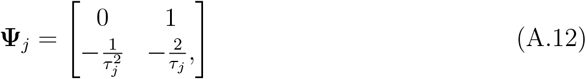

where *j* = 1, …, *N* indexes inter-population connections.

The discrete time version **D**_*δ*_ is related to **D** by

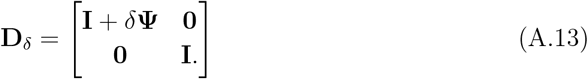

The matrix **E** has the form

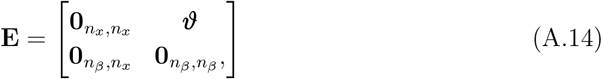

where 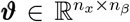 maps the connectivity to the relevant sigmoidal function and is of the form

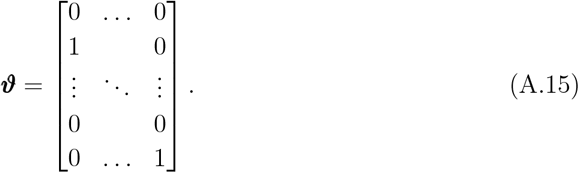

The discrete time version is

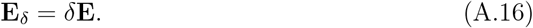

The matrix **F** is the same for both the continuous and discrete version of the model.

It has a block diagonal structure where

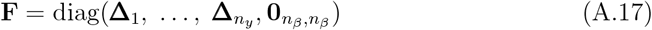

and **Δ** defines the connectivity structure of each single region model. It is a matrix of zeros or ones that specifies all the connections between the cell population types.

The matrix **G** has the form

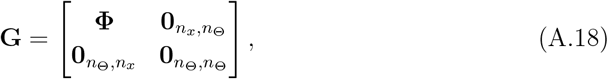

where 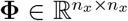 and has the form 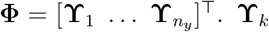 mapps the inter-regional connectivity weights to the relevant sigmoidal function and is of the form

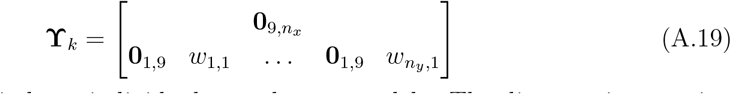

where *k* = 1, …, *n*_*y*_ indexes individual neural mass models. The discrete time version is

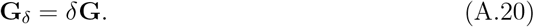

The matrix **O** is the same for both the continuous and discrete version of the model. It has a block diagonal structure where

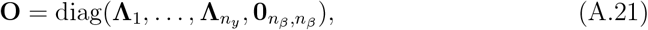

and **Λ**_*k*_ is defined to sum the relevant post-synaptic potentials to form the pyramidal membrane potential for each neural mass model then map them to the activation function as the output firing rates to other regions and is of the form

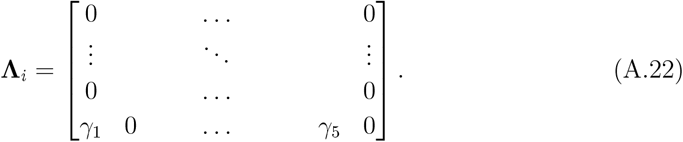

The last row of **Λ**_*i*_ indicates the post-synaptic membrane potentials that contribute to the pyramidal membrane potential.

## Appendix B. Supplementary Tables

**Table B1.** Subject data summary. The total number of seizures considered in this study, the number of days and the mean seizure rate are tabulated. Subject 5, 12, 14 were excluded in the data pre-processing phase.

| Subject | Total seizures | Number of days | Mean seizure rate (seizures/day) |
| --- | --- | --- | --- |
| 1 | 121 | 767 | 16% |
| 2 | 32 | 730 | 0.04 |
| 3 | 341 | 557 | 0.61 |
| 4 | 22 | 233 | 0.09 |
| 5 | 9 | 273 | 0.03 |
| 6 | 71 | 441 | 0.16 |
| 7 | 246 | 185 | 1.3 |
| 8 | 466 | 558 | 0.84 |
| 9 | 204 | 395 | 0.52 |
| 10 | 484 | 373 | 1.30 |
| 11 | 463 | 722 | 0.64 |
| 12 | 13 | 729 | 0.02 |
| 13 | 481 | 747 | 0.64 |
| 14 | 12 | 627 | 0.02 |
| 15 | 77 | 466 | 0.17 |

## Appendix C. Supplementary Figures

**Figure C1.**
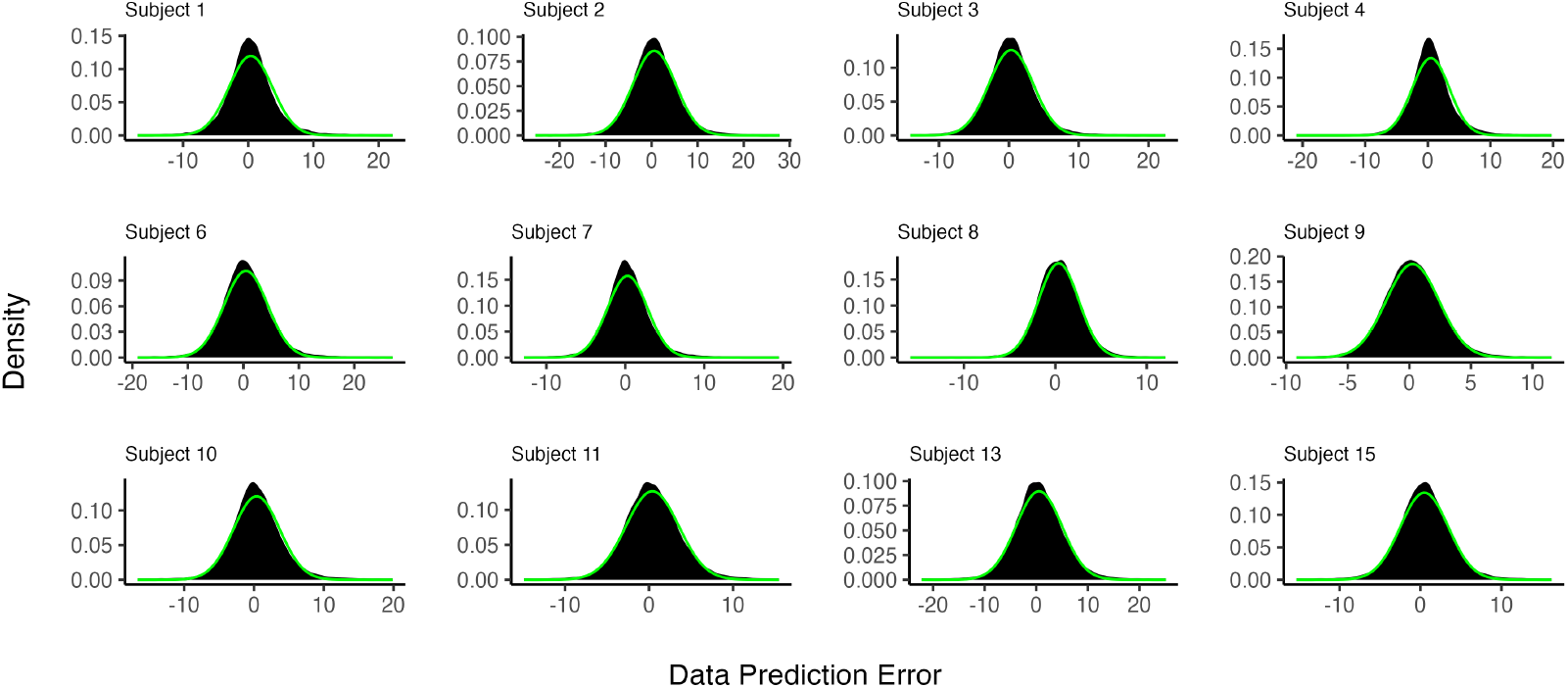
The distributions of the data prediction errors for all 12 subjects displayed as probability density functions (black histogram) plotted against normal probability density functions with the same mean and standard deviation as the data from each subject (green curves). In each plot, the y-axis represents the value of the density and the x-axis represents the data prediction errors.

